# Vaccine Preventable Disease Seroprevalence In a Nationwide Assessment of Timor-Leste (VASINA-TL) - study protocol for a population-representative cross-sectional serosurvey

**DOI:** 10.1101/2022.12.23.22283897

**Authors:** Paul Arkell, Sarah L Sheridan, Nelson Martins, Maria Y Tanesi, Nelia Gomes, Salvador Amaral, Tessa Oakley, Vanessa Solano, Michael David, Anthony DK Draper, Nevio Sarmento, Endang da Silva, Lucsendar Alves, Carlito Freitas, Filipe de Neri Machado, Celia A Gusmão, Ismael da Costa Barreto, Nicholas SS Fancourt, Kristine Macartney, Jennifer Yan, Joshua R Francis

## Abstract

**Introduction:** Historic disruption in health infrastructure combined with data from a recent vaccine coverage survey suggests there are likely significant immunity gaps to vaccine preventable diseases and high risk of outbreaks in Timor-Leste. Community-based serological surveillance is an important tool to augment understanding of population-level immunity achieved through vaccine coverage and/or derived from prior infection.

**Methods and analysis:** This national population-representative serosurvey will take a three-stage cluster sample and aims to include 5600 individuals above one year of age. Serum samples will be collected by phlebotomy and analysed for measles immunoglobulin G (IgG), rubella IgG, severe acute respiratory syndrome coronavirus-2 anti-spike protein IgG, hepatitis B surface antibody and hepatitis B core antigen using commercially available chemiluminescent immunoassays or enzyme-linked immunosorbent assays. In addition to crude prevalence estimates and to account for differences in Timor-Leste’s age structure, we will calculate stratified age-standardised prevalence estimates, using Asia in 2013 as the standard population. Additionally, this survey will derive a national asset of serum and dried blood spot samples which can be used for further investigation of infectious disease sero-epidemiology and/or validation of existing and novel serological assays for infectious diseases.

**Ethics and dissemination:** Ethical approval has been obtained from the Research Ethics and Technical Committee of the Instituto Nacional da Saúde,Timor-Leste and the Human Research Ethics Committee of the Northern Territory Department of Health and Menzies School of Health Research, Australia. Co-designing this study with Timor-Leste Ministry-of-Health and other relevant partner organisations will allow immediate translation of findings into public health policy (which may include changes to routine immunisation service delivery and/or plans for supplementary immunisation activities).

**STRENGTHS AND LIMITATIONS OF THIS STUDY:** This project is one of very few large-scale, community-based, population-representative serosurveys to be conducted in low-middle income countries.

It will provide accurate seroprevalence estimates for multiple vaccine-preventable diseases, which will immediately inform public health policy and support an ongoing programme of vaccine research in Timor-Leste and the surrounding region.

A national asset of bio-banked serum samples will be derived, which can be used in cross-sectional and prospective studies of infectious disease epidemiology, including those which evaluate disease control interventions.

Diverse, remote communities across Timor-Leste will be visited, with primary sample analysis occurring at the National Health Laboratory in Timor-Leste. Therefore, fieldwork and laboratory-related logistical challenges will need to be overcome.

## INTRODUCTION

The Democratic Republic of Timor-Leste (Timor-Leste) achieved independence in 2002. It is a half-island nation located between Australia and Indonesia with a population of 1.3 million people. The Expanded Program on Immunisation (EPI) began *circa* 1989 when Timor-Leste was still an Indonesian province. In 1999 there was significant disruption of healthcare infrastructure, including the near cessation of routine vaccine delivery. After independence was regained in 2002, the EPI was reinstated as part of a national vaccination programme, initially with single-dose measles vaccine. Hepatitis B vaccination in infancy (three doses) was introduced by 2007. Birth dose hepatitis B vaccine was introduced in 2016, along with combined measles-rubella (MR) vaccination (two doses). Vaccination against severe acute respiratory syndrome coronavirus 2 (SARS-CoV-2) began in adults in April 2021 and children above 12 years of age in October 2021.

The most comprehensive recent assessment of routine childhood vaccination coverage in Timor-Leste was a survey undertaken in 2018 which used a combination of maternal history and vaccination card review to confirm doses of vaccines given in the first and second years of life. This study found variable uptake between different vaccines and across geographic regions, and highlighted the need for further investigation of population immunity to vaccine-preventable diseases (VPDs, see table 1).^1^

**Table 1:**
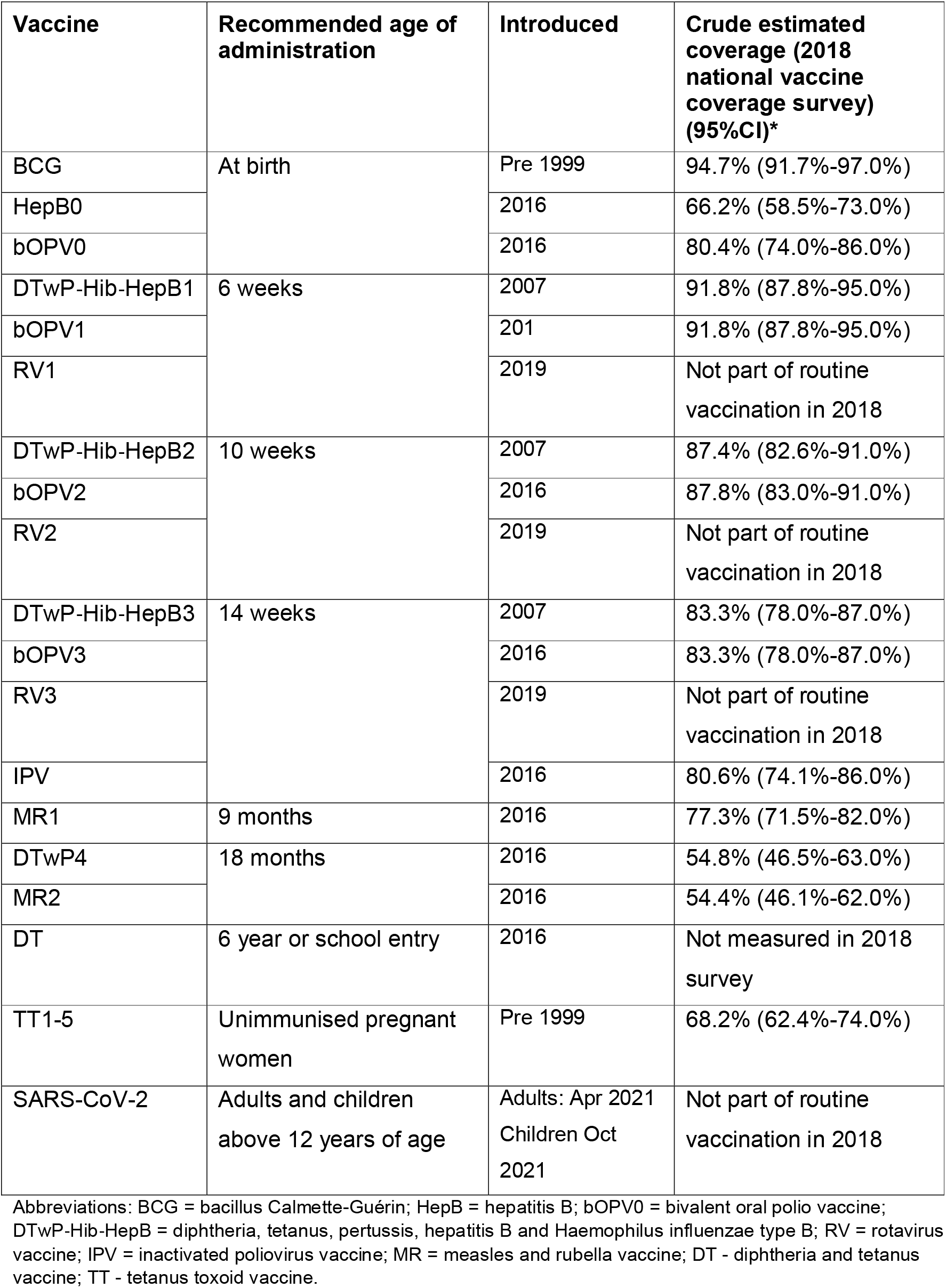
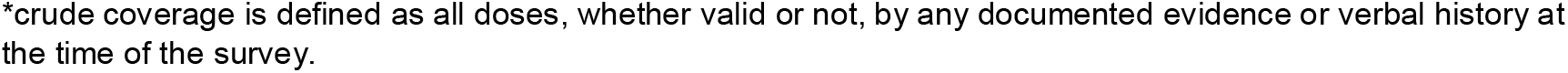
Routine Timor-Leste vaccination schedule in 2022 and estimated coverage in 2018.

| Vaccine | Recommended age of administration | Introduced | Crude estimated coverage (2018 national vaccine coverage survey) (95%CI)* |
| --- | --- | --- | --- |
| BCG | At birth | Pre 1999 | 94.7% (91.7%-97.0%) |
| HepB0 |  | 2016 | 66.2% (58.5%-73.0%) |
| bOPV0 |  | 2016 | 80.4% (74.0%-86.0%) |
| DTwP-Hib-HepB1 | 6 weeks | 2007 | 91.8% (87.8%-95.0%) |
| bOPV1 |  | 201 | 91.8% (87.8%-95.0%) |
| RV1 |  | 2019 | Not part of routine vaccination in 2018 |
| DTwP-Hib-HepB2 | 10 weeks | 2007 | 87.4% (82.6%-91.0%) |
| bOPV2 |  | 2016 | 87.8% (83.0%-91.0%) |
| RV2 |  | 2019 | Not part of routine vaccination in 2018 |
| DTwP-Hib-HepB3 | 14 weeks | 2007 | 83.3% (78.0%-87.0%) |
| bOPV3 |  | 2016 | 83.3% (78.0%-87.0%) |
| RV3 |  | 2019 | Not part of routine vaccination in 2018 |
| IPV |  | 2016 | 80.6% (74.1%-86.0%) |
| MR1 | 9 months | 2016 | 77.3% (71.5%-82.0%) |
| DTwP4 | 18 months | 2016 | 54.8% (46.5%-63.0%) |
| MR2 |  | 2016 | 54.4% (46.1%-62.0%) |
| DT | 6 year or school entry | 2016 | Not measured in 2018 survey |
| TT1-5 | Unimmunised pregnant women | Pre 1999 | 68.2% (62.4%-74.0%) |
| SARS-CoV-2 | Adults and children above 12 years of age | Adults: Apr 2021<br>Children Oct 2021 | Not part of routine vaccination in 2018 |

Abbreviations: BCG = bacillus Calmette-Guérin; HepB = hepatitis B; bOPV0 = bivalent oral polio vaccine; DTwP-Hib-HepB = diphtheria, tetanus, pertussis, hepatitis B and Haemophilus influenzae type B; RV = rotavirus vaccine; IPV = inactivated poliovirus vaccine; MR = measles and rubella vaccine; DT - diphtheria and tetanus vaccine; TT - tetanus toxoid vaccine.
\*crude coverage is defined as all doses, whether valid or not, by any documented evidence or verbal history at the time of the survey.

For many pathogens, including measles, rubella, hepatitis B and SARS-CoV-2, specific immunoglobulin G (IgG) antibodies can be detected in the blood for many years, and sometimes lifelong, following infection or vaccination. In some cases, a specific quantity and/or quality of antibody in individuals’ sera has been associated with protection from infection upon subsequent exposure. Presence above antibody cut-off levels can in some contexts infer protection, although how much such levels correlate with protection, varies on a range of factors.^2,3^ Nonetheless, community-based serological surveillance is an important tool to augment understanding of population level immunity achieved through vaccine coverage over many years and/or immunity to VPDs derived from prior infection.^4^ The results of serosurveys can be used to guide supplementary immunisation activities (SIAs), and tailor routine immunisation service delivery. There have been no previous community-based studies estimating VPD seroprevalence in Timor-Leste.

This paper describes the protocol for a first and comprehensive national population-representative serosurvey of multiple VPDs. The survey is also designed to derive a national asset of serum and dried blood spot (DBS) samples which can be used for further investigation of infectious disease sero-epidemiology in Timor-Leste.

## METHODS AND ANALYSIS

### Aim

To determine the seroprevalence of measles, rubella, hepatitis B and SARS-CoV-2 among individuals of different age-strata in Timor-Leste.

### Design

Population-representative, national cross-sectional serological survey.

### Setting

Recruitment and data collection will occur in the community (within households) across Timor-Leste. Laboratory analysis will occur at Laboratório Nacional da Saúde (LNS) in Dili, Timor-Leste.

### Sampling methods

Timor-Leste is made up of 12 municipalities and one Special Region (*Região Administrativa Especial de Oecusse Ambeno*), some of which are divided into sub-municipalities (*Posto administrativos)*. Atauro is a 14th municipality but through legacy is included as part of the Municipality of Dili. Each (sub-)municipality is divided into *sucos* (villages), which are further divided into *aldeias* (hamlets). In 2015, a national census took place in Timor-Leste. All households in the country were visited in-person, assigned a household number and global positioning (GPS) coordinates, and grouped into 2320 ‘enumeration areas’ (EAs, the boundaries of which roughly correspond to those of each aldeia, see Figure 1).

**Figure 1:**
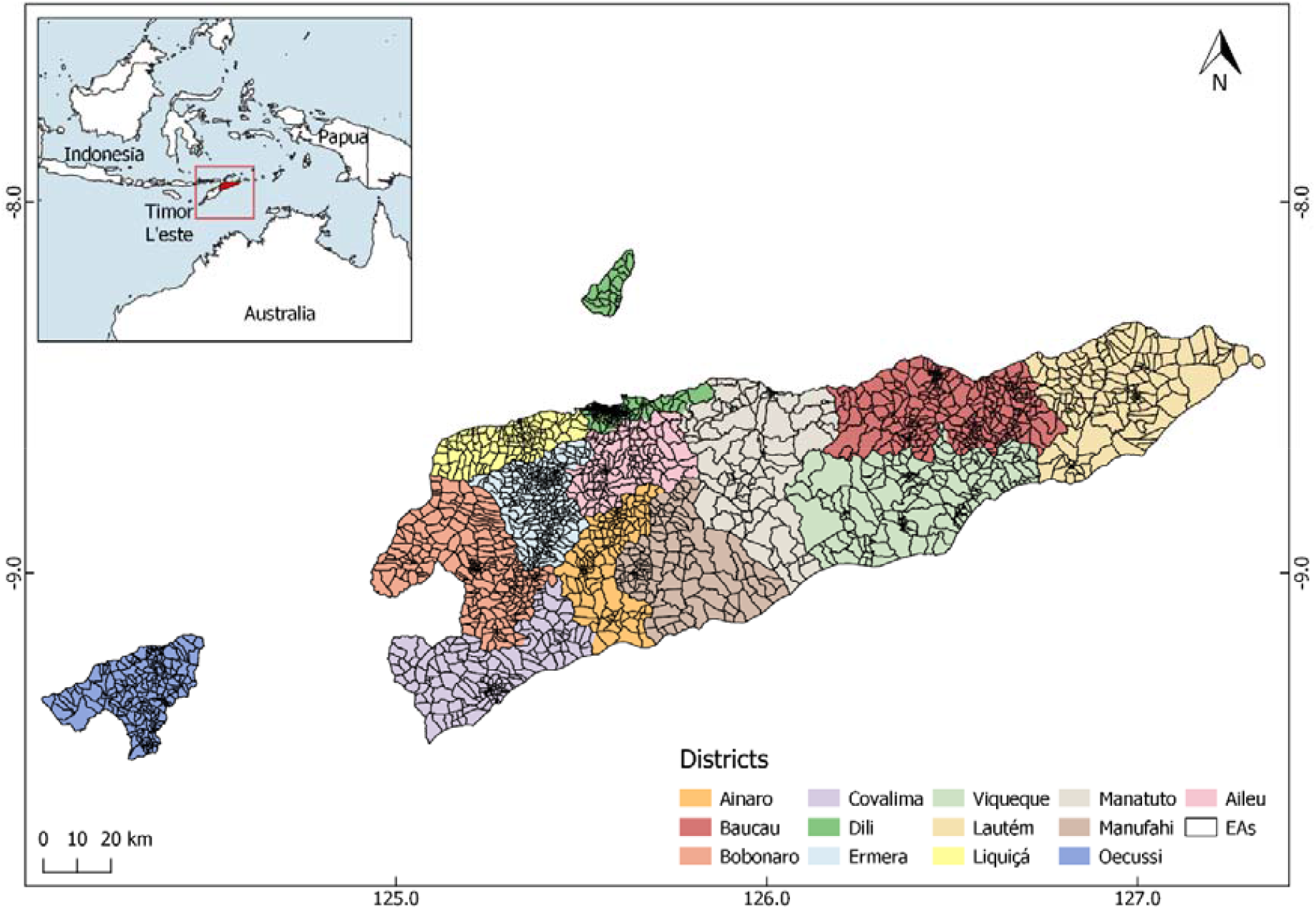
Map of Timor-Leste showing enumeration areas (EAs) within municipalities

A three-stage cluster random sample will be taken. First, a pre-specified number of EAs will be randomly selected from all EAs in the country, with probability proportionate to municipality population. Second, within each participating EA, a pre-specified number of households will be randomly selected from all households in that EA. Third, all occupants at participating households who meet eligibility criteria will be invited into the study.

A household will be defined as a dwelling unit that consists of a person or a group of related or unrelated persons, who live together in the same dwelling unit or informal shelter, who are considered as one unit and share a cooking area.

### Eligibility criteria

Household members will be eligible to participate if they are ≥1 year of age and they (or their parent/guardian) provide consent to participate in the study.

Individuals who report current illness which is compatible with coronavirus disease (COVID-19), and those who report any of the following conditions will be excluded:

- Needle phobia
- Anaemia
- Skin condition affecting phlebotomy sites
- Bleeding disorder

Additionally, individuals who cannot communicate verbally in Tetum, Portuguese or English will be excluded.

### Sample size

The sample size estimation and the proposed parameters are described in Table 2. Age groups of 1-4, 5-14, 15-24, 25-40, and >40 years have been considered as separate strata, for a range of reasons, with some including:

**Table 2:**
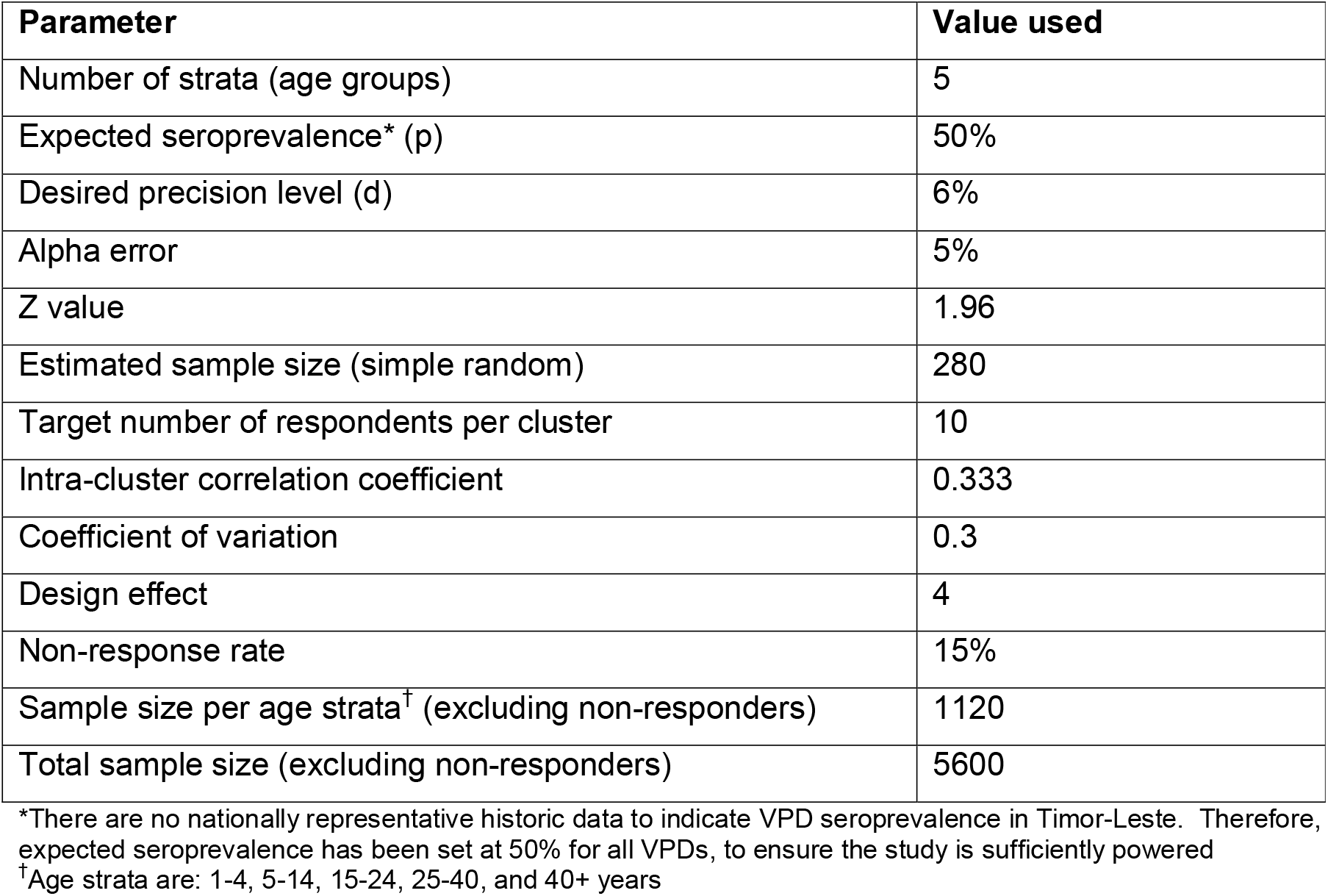
Parameters for the cluster survey sample size calculation (for national age-stratified seroprevalence estimates for all VPDs)

| Parameter | Value used |
| --- | --- |
| Number of strata (age groups) | 5 |
| Expected seroprevalence* (p) | 50% |
| Desired precision level (d) | 6% |
| Alpha error | 5% |
| Z value | 1.96 |
| Estimated sample size (simple random) | 280 |
| Target number of respondents per cluster | 10 |
| Intra-cluster correlation coefficient | 0.333 |
| Coefficient of variation | 0.3 |
| Design effect | 4 |
| Non-response rate | 15% |
| Sample size per age strata<sup>†</sup> (excluding non-responders) | 1120 |
| Total sample size (excluding non-responders) | 5600 |

\*There are no nationally representative historic data to indicate VPD seroprevalence in Timor-Leste. Therefore, expected seroprevalence has been set at 50% for all VPDs, to ensure the study is sufficiently powered
<sup>†</sup>Age strata are: 1-4, 5-14, 15-24, 25-40, and 40+ years

- Measles in children under 5 years of age because they are most likely to suffer serious sequalae from infection when compared to other age groups^5^ and between 5-14 years of age because outbreaks can occur and/or amplify in schools and other settings where groups of children from different households congregate.
- Rubella in women 15-40 years of age because future pregnancies may be at risk of congenital rubella syndrome (CRS). It is anticipated that rubella virus is circulating in Timor-Leste as there has only been recent introduction of rubella vaccination and there is very little surveillance for CRS.
- SARS-CoV-2 in children 1-12 years of age because seropositivity is likely to represent naturally acquired infection as this group are not eligible for vaccination in Timor-Leste, and so it will give an indication of the extent of local transmission which has occurred.
- Hepatitis B (surface antibody, HBsAb and core antibody, HBcAb) in children under 5 and 5-14 years of age because hepatitis B birth vaccination was introduced approximately 5 years ago and comparison of these groups will give an indication of uptake.

The required sample size for this multi-stage survey (householders within households within EAs) will need to power seroprevalence estimates with a precision of 6%. For the first step of this determination and assuming 50% seroprevalence, a simple random sample of 280 will be required for each of the five age strata. Using an intraclass correlation of 0.333, which accounts for dependence among households and households at the second and third stages, respectively, this sample size was then adjusted by the design effect of four in step 2. Thus the required effective sample size for each stratum was calculated to be 1120. As non-response is expected to be minimal, there was no need to adjust this figure by a non-response factor. Lastly, and considering the five strata, the required sample size is estimated to be 5600.

The formula for the sample size calculation is:

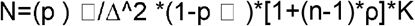

Where,

∆ = precision estimate

p □ = seroprevalence estimate

n = mean number of householders in a household estimate

ρ = intraclass correlation coefficient estimate K = number of age strata

### Fieldwork procedures

Municipalities will be visited sequentially depending on various logistical considerations including weather and road conditions, availability of staff, vehicles, and accommodation, and local municipal and health leader preference.

First, the study team leader will make a ‘coordination visit’ to the municipality during which they will explain the study procedures, receive permission to conduct the study, and discuss travel routes and gaining access to each EA with the following individuals:

- Municipality Administrator (at Municipality Office; one per municipality)
- Director of Municipality Health Service (at Municipality Health Service Office; one per municipality)
- Sub-Municipality Administrator (at Administrative Post Offices; one for each sub-municipality being visited)
- Head of Community Health Centre (at Community Health Centre Office; one for each sub-municipality being visited)
- Chief of Suco (at Suco Office;1 for every suco being visited)
- Commander of Police in Suco (at Suco Police Station; one per suco being visited, at the discretion of the Chief of Suco)
- Chief of Aldeia (at Aldeia Office; one for every aldeia being visited)

If any of these individuals are not available in-person during the coordination visit attempts will be made to contact them by telephone or through WhatsApp.

Secondly, a ‘study visit’ will be made by a whole study team, consisting of a team leader (usually non-clinical), three research nurses, and two drivers. Additionally, at the discretion of the Municipality Administrator, Sub-Municipality Administrator and/or Head of Community Health Centre, one or two local government representatives and/or one or two Community Health Centre representatives may join the study visits. It is anticipated that these individuals will primarily observe study procedures. Any involvement in participant recruitment, data collection or sample collection will be directly supervised by the appropriate study team member.

### Navigation and maps

Selected households will be identified using electronic tablets which will have GPS capability and Google Earth® software installed. Keyhole Markup Language (KML) files with GPS coordinates for all selected households in each EA will be pre-loaded onto the tablets, such that they can be used without mobile/internet connectivity. KML files are generated in QGIS. Each household location is verified using a Google Satellite base map. Study teams will also carry printed colour copies of bespoke maps for each EA, which will show the location of households in relation to roads, paths, and landmarks. These will be produced using ArcMap™ 10.4.1 and will include ESRI imagery base map, showing the location of main roads and the selected households coded by letters. The standard operating procedure (SOP) for location of households is shown in appendix 1.

### Data collection

Study teams will approach the household occupants and introduce themselves. The occupants will be asked to identify an (acting) head-of-household. If this individual is not available (or if no occupants are present), the study team will arrange to return twice to the household, and at least once on a separate day until a head-of-household is present.

Data will be collected using structured interview-questionnaires. Reponses will be entered into electronic tablets which will have REDCap® installed. This is a secure web platform for building and managing online databases which allows offline data entry^6^. Questionnaires data will be uploaded to the REDCap® secure server hosted at Menzies School of Health Research, Charles Darwin University, Darwin, Australia.

Three bespoke data collection tools have been developed (see appendices 2-4):

- Housenold questionnaire. This will be completed first. Demographic data on all household occupants (whether they are present at the time or not), will be collected by interviewing the head-of-household.
- Participant questionnaire. This will be completed if/when any household occupants agree to participate. Each will be assigned a unique identification number (participant ID number) and relevant demographic, clinical and vaccine-related data will be collected. Participants will not be asked to provide written documentation of vaccines received because a low proportion of participants in the recent vaccine coverage survey had retained this.^1^
- Unable to complete questionnaire. This will only be completed if the household questionnaire cannot be completed (i.e. if a head-of-household was not present or not willing to provide demographic data after 3 household visits). The reason for non-completion will be recorded in free-text.

### Sample collection and handling

Research nurses with training and experience in adult and paediatric phlebotomy will collect primary blood samples using appropriate infection prevention control procedures and safe management of sharps. Participants >5 years of age will undergo venepuncture using either a standard hypodermic or a winged butterfly needle with a syringe attached. Venous blood will then be injected directly into a gel serum separator tube (SST). Participants between 1-5 years of age (and those who do not consent to venepuncture but provide consent for a finger prick) may undergo capillary blood sampling through finger prick technique, in which case drops of blood will be applied directly to a paediatric gel SST. The method used will be determined on a case-by-case basis by the research nurse in the field. Table 3 shows sample volumes and collection techniques for participants in different age groups.

**Table 3:**
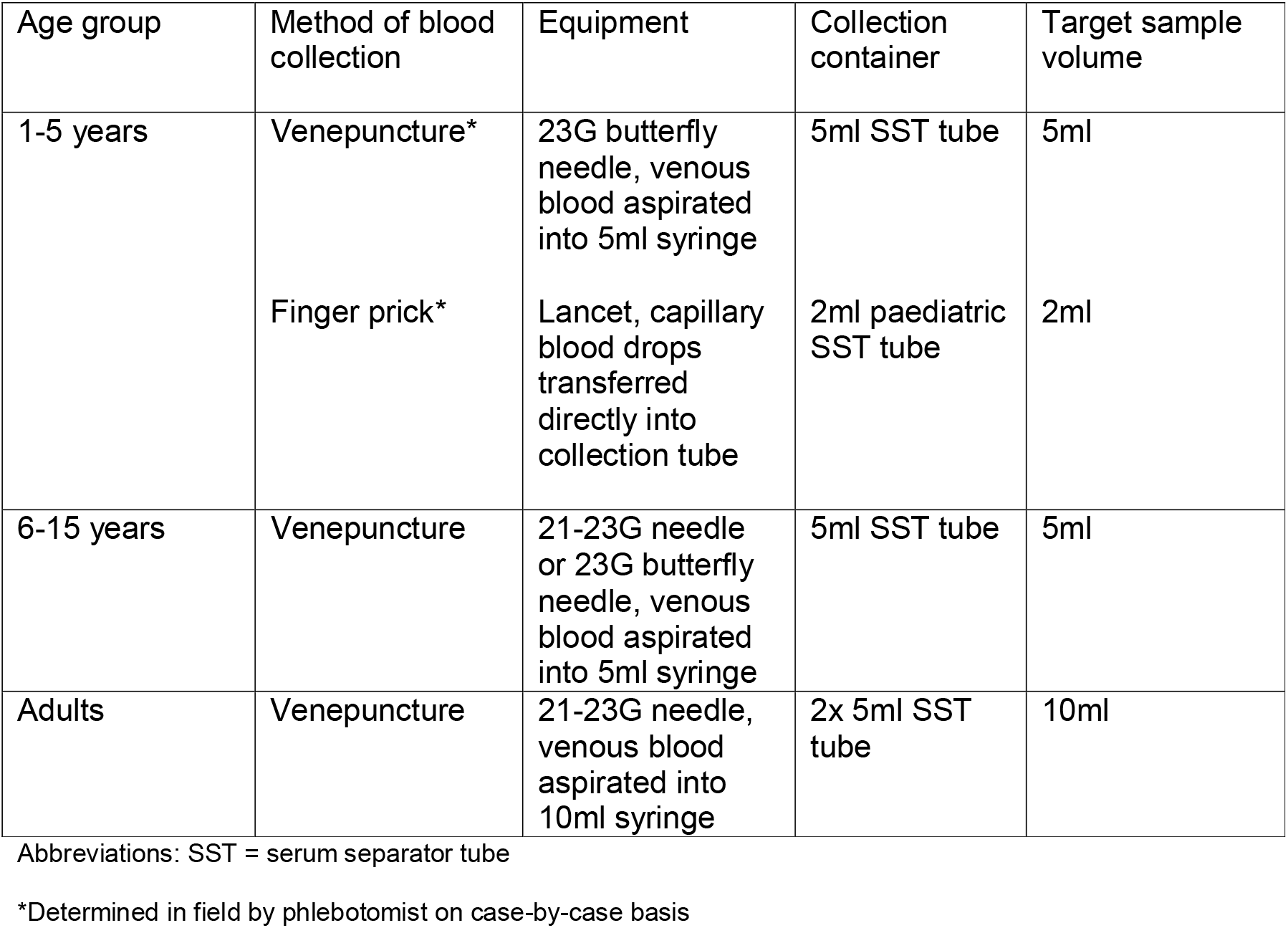
Sample volumes and collection techniques for primary sample collection by age group.

Primary blood samples will be kept at ambient temperature out of direct sunlight and allowed to clot for a maximum of eight hours (i.e. one day of fieldwork). They will then undergo centrifugation at 1,500 RCF for ten minutes and the resulting separated serum samples will be kept at 4 degrees Celsius using a portable refrigerator with battery power backup. They will be transported to LNS within five days of sample collection and will undergo primary serological analysis within two weeks of sample collection.

A secondary dried blood spot (DBS) sample will be created: For participants who undergo venepuncture, the last 300-500μL of venous blood in the syringe will be injected onto Whatman 903 filter paper marked with three 12mm diameter circles. For participants who undergo finger-prick, additional drops of capillary blood will be applied directly from the finger to the filter paper. Once the circles are saturated with blood (typically using 100-150μL blood for each circle), the filter paper will be dried at ambient temperature out of direct sunlight for four hours, then placed alongside a desiccant sachet into a plastic zip lock bag. The SOP for data and sample collection is shown in appendix 5.

### Sample analysis

Primary (serum) samples from all participants will be tested at LNS for rubella IgG (quantitative; considered positive if >10 IU/mL), SARS-CoV-2 anti-spike IgG (qualitative), hepatitis B core antibody (HBcAb, qualitative) and hepatitis B surface antibody (HBsAb, quantitative, considered positive if >10 mIU/mL) using Ortho Clinic Diagnostics® chemiluminescent assays on the Vitros ECiQ® platform, and for measles IgG using the Eurimmun® ELISA assay (quantitative, positive if >120IU/L). For quantitative assays, serological cut-offs which have been most commonly shown to correlate with protection from infection and/or those which are conventionally used in serosurveys and/or assessment of immunity have been chosen.^7–9^ Where data are somewhat conflicting, there is lack of consensus supporting a correlate-of-protection, or considerable inter-assay variability in quantitative determination has been observed, secondary (exploratory) analyses using alternative cut-offs may also be undertaken (for example 200IU/L and/or 250IU/L for measles IgG).^3^

Additionally, samples from participants residing in Dili Municipality (excluding Atauro) will be tested for hepatitis B surface antigen (HBsAg, qualitative). This marker denotes active hepatitis B infection which may have significant health implications and will therefore only be tested in Dili municipality where there is a hepatology clinic in which participants can receive further assessment and follow-up. Any samples which are positive for HBsAg will also be tested for hepatitis B envelope antigen (HBeAg, qualitative) and hepatitis B envelope antibody (HBeAb, qualitative) testing using Ortho Clinic Diagnostics chemiluminescent assays on the Vitros ECiQ platform, and hepatitis B viral load (HBVL, quantitative) using the Cepheid® assay on the GeneXpert platform. All testing will be carried out according to manufacturers’ instructions and cited serological cut-off values. For qualitative assays, samples with borderline/indeterminate results will be considered negative, apart from HBsAg where the test will be repeated and both results reviewed alongside other hepatitis B results by an appropriately qualified clinical member of the research team who will decide whether the participant should be referred to the hepatology clinic for repeat sampling and clinical assessment.

Assays have been chosen based on their previous performance in seroprevalence studies, immediate availability for shipment to Timor-Leste, and local laboratory expertise in operating these types of assays (with ongoing capacity building for serological testing in LNS). The measles IgG assay is quantitative and calibrated against a World Health Organisation (WHO) Standard (NIBSC, Anti-Measles serum, 3rd International Standard 97/648). It showed acceptable performance when assessed in a recent study of concordance between commercially available assays (concordance for samples with positive/negative status = 90%/100%)^10^. The rubella IgG assay is quantitative and calibrated against a Centers for Disease Control and Prevention (CDC) standard (Low Titer Rubella Standard) and a World Health Organization (WHO) standard (1st International Rubella IgG Standard). While concerns around standardisation of rubella IgG assays are noted^11^, this assay showed acceptable performance when assessed in a recent study of concordance between automated immunoassays (concordance for samples with negative/positive status = 90.6%/91.1%)^12^. The hepatitis B surface antibody assay is quantitative and showed high sensitivity (97.1%) and specificity (97.9%) when evaluated in a panel of sera from healthcare workers and patients^13^. The SARS-CoV-2 anti-spike IgG assay has high sensitivity (93.3%, >21 days post infection) and specificity (100%), which compares favourably to many other available immunoassays^14^, and has been used in several serological surveillance studies^15–17^.

### Provision of results to participants

The majority of testing in this study will be for antibodies against VPDs (either IgG or total antibody). Results will therefore only indicate whether an individual has been previously infected and/or vaccinated against each disease at some time in the past and will not provide information on current infection. While seronegative participants may be at risk of future infections (and may benefit from vaccination), individual notification of results and provision of vaccines to all seronegative study participants is not considered feasible in this large cross-sectional study. Instead, all participants will be advised of the benefits of routine vaccination and immunisation clinics in their area, as well as on any forthcoming SIAs which may occur as a result of this study.

In addition to antibody tests, serum from participants within Dili Municipality (excluding Atauro) will be tested for HBsAg. This marker denotes active hepatitis B infection which may have significant health implications and will therefore only be tested in Dili municipality where there is a hepatology clinic in which participants can be seen. Participants who test positive for HBsAg will be contacted by telephone to discuss their results and will be offered assessment including biochemical and radiological investigation of liver function and consideration of antiviral treatment in-line with international clinical guidelines^18^. This approach has been successful and has been acceptable to participants in a smaller serological surveillance study including hepatitis B testing among healthcare workers in Timor-Leste^19^.

### Sample storage

Primary (serum) samples will be stored at -80 degrees Celsius and secondary (DBS) samples will be stored at 4 degrees Celsius at LNS for 10 years. These may undergo additional serological analyses to further investigate infectious disease sero-epidemiology in Timor-Leste and/or validating existing and novel serological assays for infectious diseases, pending successful funding application and appropriate ethical approval.

### Fieldworker training

Field workers will undergo one week of formal in-person training in study procedures. Days 1-2 will be classroom based and will include sessions on ‘the study protocol’, ‘field team composition’ (structure, members, responsibilities), ‘logistics, technology and map reading’, ‘recruitment and consent’, and ‘collection of data using interview questionnaires’. Days 3-5 will be practical and will include demonstrations and training in adult and paediatric phlebotomy and finger-prick techniques, infection prevention control procedures, and the use of personal protective equipment (PPE). These skills will be assessed formatively throughout the training and summatively using pre- and post-session assessments. Training will be delivered by PA, JF, NSSF and/or JY who are clinicians with experience of epidemiological and clinical research in Timor-Leste.

### Laboratory team training

Laboratory training will occur in the Serology Department of LNS. Training will be delivered by PA and TO who have significant experience with ELISA and chemiluminescent techniques, including in Timor-Leste. The focus of training will be on assay verification and quality assurance, as well as procedures for sample processing, analysis and storage.

### Data storage and handling

Field data will be stored in the REDCap® secure server hosted at Menzies School of Health Research, Charles Darwin University, Darwin, Australia, until analysis. Laboratory data (i.e. serology worksheets and results) will be stored on the password-protected LNS laboratory information system (SchuyLab®) until analysis. Deidentified field and laboratory datasets will be downloaded and stored as password-protected databases on computer(s) at Menzies School of Health Research, Timor-Leste Office, Dili, Timor-Leste, where they will be linked using participant ID numbers and analysed. Only named investigators who are working directly on this project will have access to data.

### Statistical analysis plan

Primary data analysis will occur at the end of the study, once all fieldwork is complete and all samples have been analysed. Interim analyses may also occur upon reasonable request from the Timor-Leste MoH or other partner organisation. As a multi-stage sampling survey design will be used to select participants, sampling weights will be calculated at each stage. These weights will reflect a participant’s inverse probability of selection at a particular stage, be it at the EA level, the household level or the householder level. Furthermore, these weights will subject to both non-response adjustment and finite population correction. Measures of prevalence will be age-standardised using the standard population for Asia given by the International Network for the Demographic PNGIMR 2018, Papua New Guinea MIS 2016-2017.^20^ To account for this design, the ‘svy’ data commands in Stata (version 16, StataCorp, College Station, TX, USA) will be used for ally analyses.

Characteristics of participants will be summarised using weighted descriptive statistics. Frequencies and proportions will be used to describe categorical distributions, whilst means and standard deviations will be used to describe continuous variables. In the presence of non-normality, medians and interquartile ranges will be reported. Univariable and multivariable binary logistic regression will be undertaken to model age, the independent variables of primary interest with the five VPD outcomes. In addition to this variable, other variables known to be risk factors of VPD, such as sex and travel history, will be subjected to a manual backward stepwise procedure. Variables with a p-value ≥ 0.20 will not be retained. The Hosmer-Lemeshow test will be used to test the goodness of fit of each multivariable model. A p-value < 0.05 will be considered statistically significant with Odds Ratios (OR), 95% confidence intervals (CI) and p-values calculated for age and sex.

## Data Availability

Study protocol paper, therefore no data currently

## ETHICS AND DISSEMINATION

### Informed consent

Each prospective participant will receive a participant information sheet which will be printed in English and Tetum. They will also be provided with a verbal explanation of the study rationale and procedures. This will include potential risks and benefits of sample collection, specific tests which their sample will undergo, the fact that they will not receive notification of any results (with the exception of a positive HBsAg, tested in Dili Municipality only) and the possibility that their sample will undergo additional analyses for evidence of communicable diseases during the next 10 years. They will be given up to 30 minutes to ask questions and decide whether they wish to participate, and will then provide informed, written consent by signing a consent form. For individuals under 16 years of age, verbal assent will be sought, in addition to written consent from their parent or guardian. This study has received ethical approval from the Research Ethics and Technical Committee of the Instituto Nacional da Saúde,Timor-Leste (Reference: 875 MS-INS/DGE/IX/2021) and the Human Research Ethics Committee of the Northern Territory Department of Health and Menzies School of Health Research, Australia (Reference: 2021-4064).

### Protocol amendments

Any modifications to the protocol which may impact on the conduct of the study will be documented in a formal protocol amendment and approved by both Research Ethics Committees prior to implementation of the changes. The Research Ethics Committees will also be notified of any minor corrections/clarifications or administrative changes to the protocol, which will be documented in a protocol amendment letter.

### Adverse events

Data on adverse events will be collected throughout the study, with participants (or their parent/guardian) being informed of the risks of phlebotomy (including bruising, bleeding and infection), how to recognise these, and how to contact the study team if they occur. Adverse events will be reported to the Principal Investigator. In cases where infection or any other serious adverse event has occurred, the Principle Investigator will conduct a review of the study visit and decide whether any phlebotomy retraining or change in practice is required and/or whether recruitment to the study should be paused.

### Strengths and limitations

This project will produce accurate, nationally representative seroprevalence data for multiple VPDs and relevant age-groups, which has not been achieved in Timor-Leste previously. It has been co-designed by investigators at Menzies School of Health Research (Timor-Leste Office), the National Centre for Immunisation Research and Surveillance (Australia), the MoH (Timor-Leste), LNS (Timor-Leste), and the WHO (Timor-Leste Office) according to local research and public health priorities. This will allow immediate translation of findings into public health policy (including potentially changes to routine immunisation service delivery and/or plans for SIAs). Additionally, the survey will derive a national asset of serum and dried blood spot (DBS) samples which can be used for further investigation of infectious disease sero-epidemiology in Timor-Leste and/or validating existing and novel serological assays for infectious diseases.^21–25^ Engagement with local administrative and health leaders and maximisation of participant choice and welfare have been central to the design, including ensuring all individuals diagnosed with active hepatitis B during the study have access to appropriate further investigation and follow-up.

Risks and limitations include the ongoing global outbreak of SARS-CoV-2 (which may delay/prohibit study visits), disruption of supply of field and laboratory consumables to Timor-Leste (which may delay/increase the cost of laboratory analysis), natural disasters such as flooding, and potential unwillingness of individuals to participate in provision of data and/or samples (which may affect recruitment, potentially disproportionately among children).

### Dissemination / knowledge transition plan

After each interim analyses, results will be shared with Timor-Leste MoH partners in the form of an oral presentation and in a written report. Following completion of the study, results will be shared with Timor-Leste MoH, other partner organisations, and local administrative and health leaders for EAs where the study took place (Municipality Administrators, Directors of Municipality Health Services, Sub-Municipality Administrators, Heads of Community Health Centres, Chiefs of Sucos, Chief of Aldeia), in the form of a written report. Results will also be submitted for publication in peer-reviewed journals and presented at relevant international conferences.

## CONCLUSION

Historic disruption in health infrastructure including to the delivery of vaccines combined with data from a recent vaccine coverage survey suggests there are likely significant immunity gaps to VPDs and high risk of outbreaks in Timor-Leste. Targeted seroprevalence studies including healthcare workers in Timor-Leste have identified lower than expected seropositivity against measles, high seropositivity against SARS-CoV-2 and a high prevalence of active hepatitis B infection.^19,26^ This study will fill a crucial gap in understanding of national population immunity against VPDs and will guide routine immunisation service delivery, plans for SIAs, and an ongoing programme of vaccine research in Timor-Leste.

## AUTHOR CONTRIBUTIONS

PA, SLS, NM, SA, NS, F, FNM, NSSF, KM, JY and JRF conceived the study. PA, SLS, NM, MYT, SA, ADKD, NS, LA, CF, FNM, CAG reviewed existing literature and performed situation analysis. PA, MYT, SA, VS, LA, CAG, ICB determined the fieldwork procedures and designed data collection tools. PA, NG, TO, NS, ES, LA, ICB determined laboratory procedures. PA, MD, ADKD, NS, NSSF drafted the data and statistical analysis plan. NM, MYT, SA, NS, CF, FNM, CAG planned community engagement, obtained ethical approval and lead other regulatory aspects of the study. All authors reviewed and commented on the final manuscript.

## COMPETING INTERESTS

All authors declare no competing interests for this study.

Appendix 1: Standard operating procedure for location of households

Appendix 2: Household questionnaire

Appendix 3: Participant questionnaire

Appendix 4: Did not complete questionnaire

Appendix 5: Standard operating procedure for data and sample collection

